# A hospital-based study to assess various biomarkers for prognostic prediction of clinical outcome in COVID-19 disease

**DOI:** 10.1101/2022.05.25.22275583

**Authors:** Abhishek Padhi, Dipika Shaw, Shagufta Khatoon, Swekcha Ranjan, Anudita Bhargava, Sanjay Singh Negi

## Abstract

COVID-19 pandemic has inflicted a painful unforgettable number of deaths throughout the world. Hematological inflammatory and organ-specific biomarkers are universally practiced in helping clinical decisions in various infectious diseases. Accordingly, their role in predicting progression and severity, and fatal outcome of COVID-19 was investigated to take initial appropriate treatment measures to reduce associated mortality.

**Methods:** The retrospective analysis of a total of 126 COVID-19 cases representing mild, moderate, severe, and succumbed cases were assessed for the pattern of hematological, inflammatory, and organ-specific biomarkers.

**Results:** A total of 126 proven cases of SARS-CoV-2 infection were retrospectively analyzed for the association of various biomarkers with the COVID-19 disease progression. The CBC analysis showed that the median TLC was high for the severe group of both males (12.49 × 10^3^/μl) and females (14.23 x10^3^/μl). Similarly, the neutrophil count was also found high in the severe group, whereas the monocytes count showed low median values in severe cases, but both these parameters had no significant difference among the males and the females. The platelet count showed a significant difference (p=0.018) among the non-severe and severe groups between males and females. Among inflammatory markers, D-dimer, CRP, LDH, and APTT showed a higher median value in severe cases among both the males and females while ESR value was higher in non-severe cases and ferritin showed similar values in both severe and non-severe cases. The liver and kidney function parameters were also analyzed and a significant P-value was found for ALP (p=0.004), ALT (p=0.032), and AST (p=0.009) in the non-severe vs. severe category of COVID-19 patients.

**Discussion:** High TLC, neutrophilia, lymphopenia, thrombocytopenia, and eosinopenia are the potential risk factor for the progression of COVID-19 disease for severe and fatal outcomes. Inflammatory markers of D-dimer, CRP, LDH, APTT, and ferritin above normal range also carries the potential risk of severe and fatal outcome in COVID-19 disease. Higher ALT, AST, and serum creatinine may also carry a poor prognosis.

## Introduction

In December 2019, the first case of COVID-19 clinically presented with atypical pneumonia was reported from China (Wuhan) (1). Uncertainly throughout the world, the alarming rise in COVID-19 h a s been encountered. On 11^th^ March 2020, World Health Organization (WHO) declared COVID-19 is a global pandemic that devastated humanity worldwide and inflicted an unprecedented dramatic loss of millions of lives (2). As of 20^th^ May 2022, globally, the cumulative confirmed cases were 521,920,560, including 6,274,323 deaths (3).

These figures were 43,131,822 for confirmed cases in India, including 524,323 deaths. (3) COVID-19 pandemic has continuously shown its catastrophic effect as morbidity and mortality are still being reported from various affected countries, including India.

Based on the COVID-19 treatment guideline, the disease was clinically categorized from asymptomatic to mild/moderate, and severe presentation poses a challenge to understand its progression in the patients and better preparedness to prevent worsening of COVID-19 infection succumbed cases (4). Various literature suggests that host cellular factors and virus components were involved in the pathogenesis of the COVID-19 disease, leading to the up-the surge of the various biological parameters indicating the level of tissue damage or infection (1,5,6).

Thus, understanding the different diagnostic biomarkers might predict COVID-19 disease severity which may play a crucial role in the effective and appropriate therapeutic management of the infected cases before their progression reach a severe and subsequent fatal stage (7,8) Various hematological, biochemical, inflammatory, and immunological biomarkers have been studied and reported from across the globe for a better understanding of COVID-19 pathogenesis to identify, stratify, and predict their prognostic efficacy in guiding the better medical management (9). Since these laboratory biomarkers are cost-effective, they may have the potential preferred modality existing before us to provide the early indication of deciding and devising the treatment management strategies to prevent the possible further progression of mild/moderate into severe and fatal stage of highly unpredictable COVID-19 disease (9,10).

Several published literature has shown an elevated level of white cell count (WBC), creatinine, liver function test (LFT), kidney function test (KFT) markers, C-reactive protein (CRP), D-dimer, erythrocyte sedimentation rate (ESR), ferritin, interleukin-6 (IL-6), neutrophils whereas the decreased level of lymphocytes and thrombocytes were associated with severe and fatal cases of COVID-19(11–15). However, the various factors like geographical variation and associated immune response emergence of multiple mutants of SARS-CoV-2 restrict these studies’ findings from being generalized for the prognostic indication (16–18). Moreover, there is no published data from central India on biomarkers during different clinical stages of COVID-19 disease.

Therefore, to facilitate risk stratification of COVID-19 disease, this study has been undertaken retrospectively on COVID-19 positive patients admitted to All India Institute of Medical Sciences (AIIMS), Raipur, during the second wave of the pandemic to assess the laboratory-derived complete blood count (CBC), KFT, LFT, and inflammatory biomarkers during the different clinical manifestations of COVID-19 disease in mild/moderate, severe, and fatal clinical outcome in COVID-19 infected male and female patients from Raipur, Chhattisgarh, a the central state of India.

## Material and methods

Retrospectively, a total of 126 proven cases [positive for the presence of SARS-CoV-2 virus in their clinical specimen of nasopharyngeal and oropharyngeal swabs by real-time polymerase chain reaction (RT-PCR)] of COVID-19 patients who were admitted to AIIMS Raipur between March to June 2021 during the second wave of the COVID-19 pandemic in India were included in the present study. According to the criteria and advisory issued by the Ministry of Health & Family Welfare, the patients were categorized into two groups such as non-severe(mild and moderate) and severe (critical and death) (19–21). Severe patients were admitted to the Intensive care unit (ICU) with oxygen support, and non-severe patients were kept in the isolation ward with regular observation and medication.

All the patient’s laboratory findings include complete blood count (CBC), inflammatory markers [D-Dimer, ESR, CRP, lactate dehydrogenase (LDH), ferritin, and activated partial thromboplastin time (aPTT)], liver function test (LFT), and kidney function test (KFT) were collected. Hematological tests were performed using the Mindray BC-6800 system through the electrical impedance method. D-dimer was detected using Enzyme-Linked Immunosorbent Assay (ELISA). CRP value was obtained using the electrochemicalilumisence method on Cobas 702 Autoanalyzer.

Outcome analyses for different biomarkers were performed in the COVID-19 mild, moderate, critical, and death categories associated with females and males. Further, with the total number of enrolled cases (n=126), the combined category [non-severe (mild+ moderate) and severe (critical+ death)] analysis was performed.

### Ethics

The present study was approved by the Institutional Ethical Committee (IEC), All India Institute of Medical Science (AIIMS), Raipur on IEC number AIIMSRPR/IEC/2021/705.

### Statistical analysis

Descriptive analysis, including normality tests, was carried out for all variables using Jamovi (version 1.6.23.0). Shapiro–Wilk test was performed to analyze the Gaussian distribution of the dataset. Median was used to depict as the data did not normally distribute. To evaluate the a significant difference between the groups Mann–Whitney-U test (not normal distributed) and the Student t-tests (normal distributed) were used. Box plots were constructed using GraphPad Prism (Version 7) for each biological parameter.

## Results

Among the total enrolled proven COVID-19 patients (n=126) cases, 46 cases were female and 80 were male. Among the total cases (n=126), fifty patients (thirty-four males and sixteen females) unfortunately death to COVID-19 infection during their treatment. From the remaining cases (n=76), twenty patients were categorized in the mild category, including female (n=10) and male (n=10), twenty-one cases belonged to the moderate category, including female (n=7) and male (n=14), and thirty-five patients belong to the critical category including female (n=13) and male (n=22). In the combined category, forty-one patients [mild (n=20) and moderate (n=21)] were designated as a non-severe group, whereas the remaining eighty-five patients [critical and death] belong to the severe.

### CBC Analysis in COVID-19 patients

#### Hemoglobin (Hb)

The median value was low, i.e., 10.80 g/dl in non-severe females, and was a little high, i.e., 11.30 g/dl in severe COVID-19 females, whereas approximately similar in non-severe and severe COVID-19 males, i.e., 13.7 and 13.1 g/dl, respectively. A statistically significant value was found between the female and males in the non-severe (p= 0.003) and severe (p<0.001) cases, whereas no significant difference was observed in the case of non-severe vs. severe (p= 0.523) COVID-19 cases (Figure 1, Table 1). The range of variation in Hb was high in dead (8.3-16 g/dl) compared to moderate (10.3-16.5 g/dl) and mild (9.1-15.5 g/dl) and low in critical (11.9-16.5 g/dl) COVID male patients. The range of variation in Hb was less in a female with mild (8.1-11.6 g/dl) symptoms compared to moderate (6.5-16.2 g/dl), critical (7.7-15.5 g/dl), and dead (7.1-15.2 g/dl) COVID-19 patients.

**Table 1:** Biological markers in severe and non-severe COVID-19 cases (Female vs. Male)

| Variable<br>(Median;IQR) | Non-severe<sup>#</sup> COVID-19 |  | P-value | Severe<sup>@</sup> COVID-19 |  | P- value |
| --- | --- | --- | --- | --- | --- | --- |
|  | Female (n=17) | Male (n=24) |  | Female (n=29) | Male (n=56) |  |
| CBC markers |  |  |  |  |  |  |
| Hb (g/dl) | 10.8 (9.8-12.1) | 13.75 (12.5-14.1) | **0.003\*** | 11.25 (9.6-12.8) | 13.1 (12.4-14.46) | **<0.001\*** |
| TLC (10<sup>3</sup>/μl) | 10 (7.42-14.04) | 12.37 (7.1-15.6) | 0.701 | 12.49 (9.3-17.48) | 14.23 (11.8-17.3) | 0.141 |
| Neutrophils (%) | 84 (69-87) | 77.51 (64.8-84.9) | 0.451 | 90 (79.7-93.4) | 91.31 (85.5-94.3) | 0.169 |
| Lymphocytes (%) | 12.5 (6.4-16) | 13.85 (10.5-16.6) | 0.771 | 6.9 (2.86-10.9) | 3.3 (2.2-9.1) | 0.114 |
| Monocytes (%) | 7.8 (5-10.2) | 5.8 (3.9-11.2) | 0.723 | 3.8 (2.2-6.3) | 3.8 (2.8-5.3) | 0.956 |
| Eosinophils (%) | 2.4 (2-2.7) | 1.9 (1.06-3.07) | 0.570 | 2.1 (0.89-2.86) | 2.72 (1.89-3.7) | **0.019\*** |
| Platelet (10<sup>3</sup>/μl) | 271 (232-287) | 259 (164.5-277.5) | **0.018\*** | 221.6 (157-304) | 284 (195-362.25) | 0.059 |
| Inflammatory markers |  |  |  |  |  |  |
| D-dimer (μg/ml) | 0.8 (0.27-1.57) | 0.54 (0.26-1.8) | 0.598 | 1.69 (1.2-2.4) | 2.81 (1.18-5.2) | **0.002\*** |
| CRP (mg/L) | 59.48 (40.5-83.1) | 27 (17.9-67.2) | 0.103 | 74.2 (45.5-89.2) | 64.01 (32.06- 81.51) | 0.435 |
| Lactate dehydrogenase | 562 (407-762) | 355 (280.5-459.5) | **0.001\*** | 601 (489-783) | 600 (474.5-715) | 0.451 |
| ESR | 67.5 (55- 95) | 74 (40.5-102) | 0.589 | 55 (40-100) | 50 (34.25- 72.7) | 0.287 |
| APTT | 31.4 (29.9-36.8) | 32.85 (28.6-39.47) | 0.642 | 39.9 (28.3-48.2) | 30.8 (26.9-38.2) | 0.498 |
| Ferritin | 524 (267-790) | 819 (490.3-995.5) | 0.535 | 512 (371.7-682) | 811.5 (664.8-1072.2) | **<0.001\*** |
| Kidney function test markers |  |  |  |  |  |  |
| Urea | 26 (22-45) | 38.5 (28.25-45.5) | 0.118 | 49 (31-77) | 53 (38.75-86) | 0.247 |
| Creatinine | 0.91 (0.68-1.1) | 1.05 (0.89-1.3) | 0.072 | 0.81 (0.66-1.26) | 0.93 (0.8-1.7) | 0.021 |
| Uric acid | 4.7 (4.3-8) | 4.95 (3.7-7.4) | 0.916 | 3.4 (2.6-4.1) | 4.4 (3.4-5.9) | **0.004\*** |
| Sodium | 140 (135-141) | 139 (136- 140.2) | 0.396 | 138 (136-142) | 139 (135.7-143) | 0.636 |
| Potassium | 3.8 (3.4-4.3) | 3.95 (3.5-4.2) | 0.614 | 4.2 (3.7-4.5) | 4.35 (4-4.9) | 0.130 |
| Chloride | 99 (97-102) | 101.5 (97.7-103.2) | 0.358 | 104 (101-106) | 103.5 (100-105) | 0.511 |
| Liver function test markers |  |  |  |  |  |  |
| Total Bilirubin | 0.66 (0.53-0.71) | 0.77 (0.62-0.91) | 0.266 | 0.64 (0.47-0.84) | 0.755 (0.51-0.98) | 0.240 |
| Direct Bilirubin | 0.2 (0.14-0.27) | 0.2 (0.17-0.25) | 0.740 | 0.17 (0.12-0.23) | 0.24 (0.4-0.31) | 0.095 |
| **Indirect Bilirubin** | 0.43 (0.36-0.5) | 0.53 (0.44-0.6) | 0.256 | 0.48 (0.31-0.6) | 0.49 (0.36-0.63) | 0.482 |
| **Alkaline Phosphatase** | 86 (66-93) | 83 (62.7-107.7) | 0.351 | 110 (96-124) | 121 (92.75-144.25) | 0.944 |
| **Total Protein** | 5.98 (5.35-6.2) | 6.7 (5.9-7.1) | **0.018\*** | 6.04 (5.7-6.79) | 6.14 (5.5- 6.7) | 0.407 |
| **Alkaline amino phosphates** | 49 (32-56) | 77 (42-104) | 0.146 | 39 (26-56) | 41 (28.7-60.2) | 0.630 |
| **Aspartate aminotransferase** | 68 (51-76) | 47 (32-74.25) | 0.365 | 40 (24-61) | 43.5 (31.2-58) | 0.797 |

#-Mild and Moderate cases; @ Severe and Death

**Figure 1:**
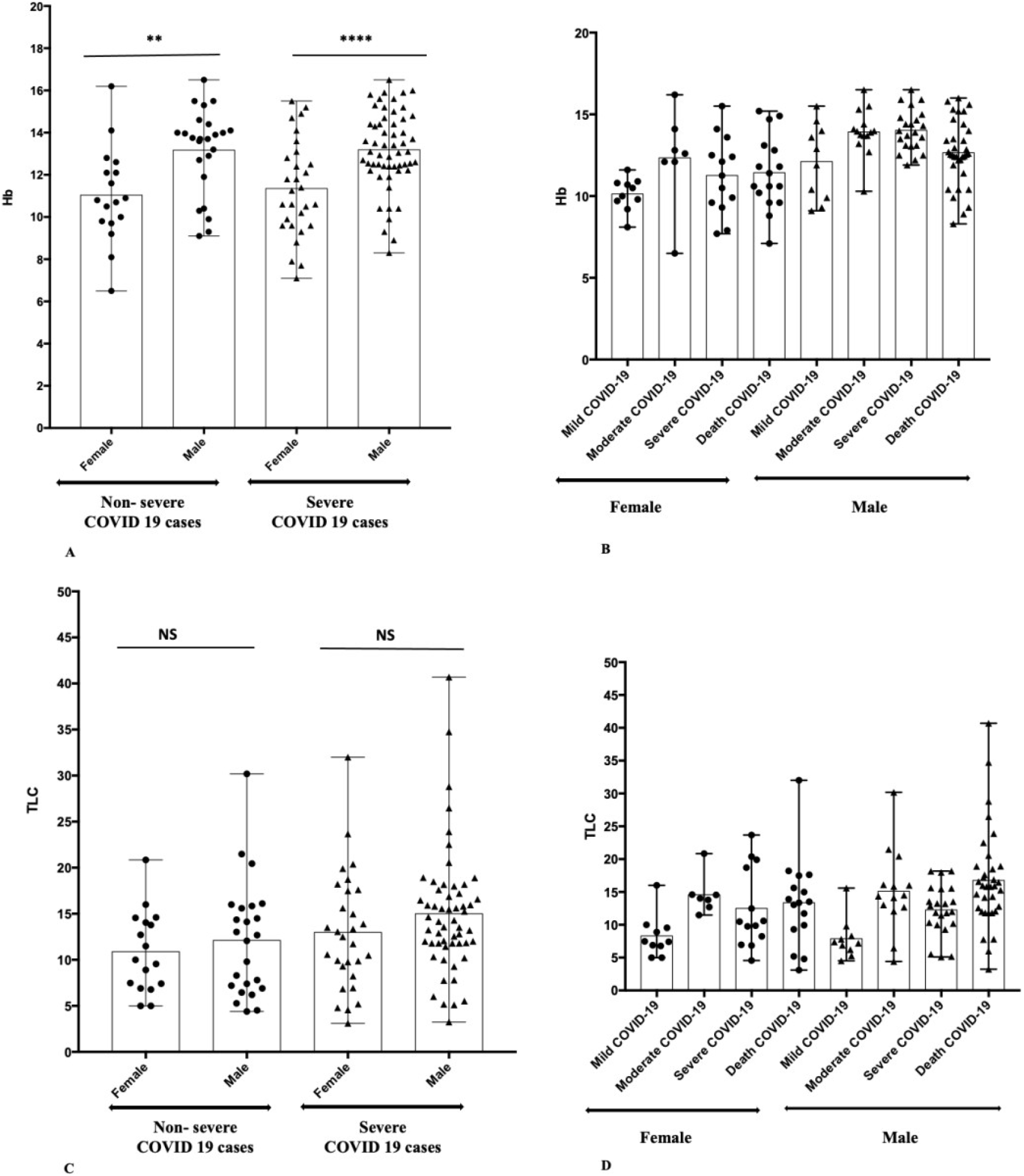
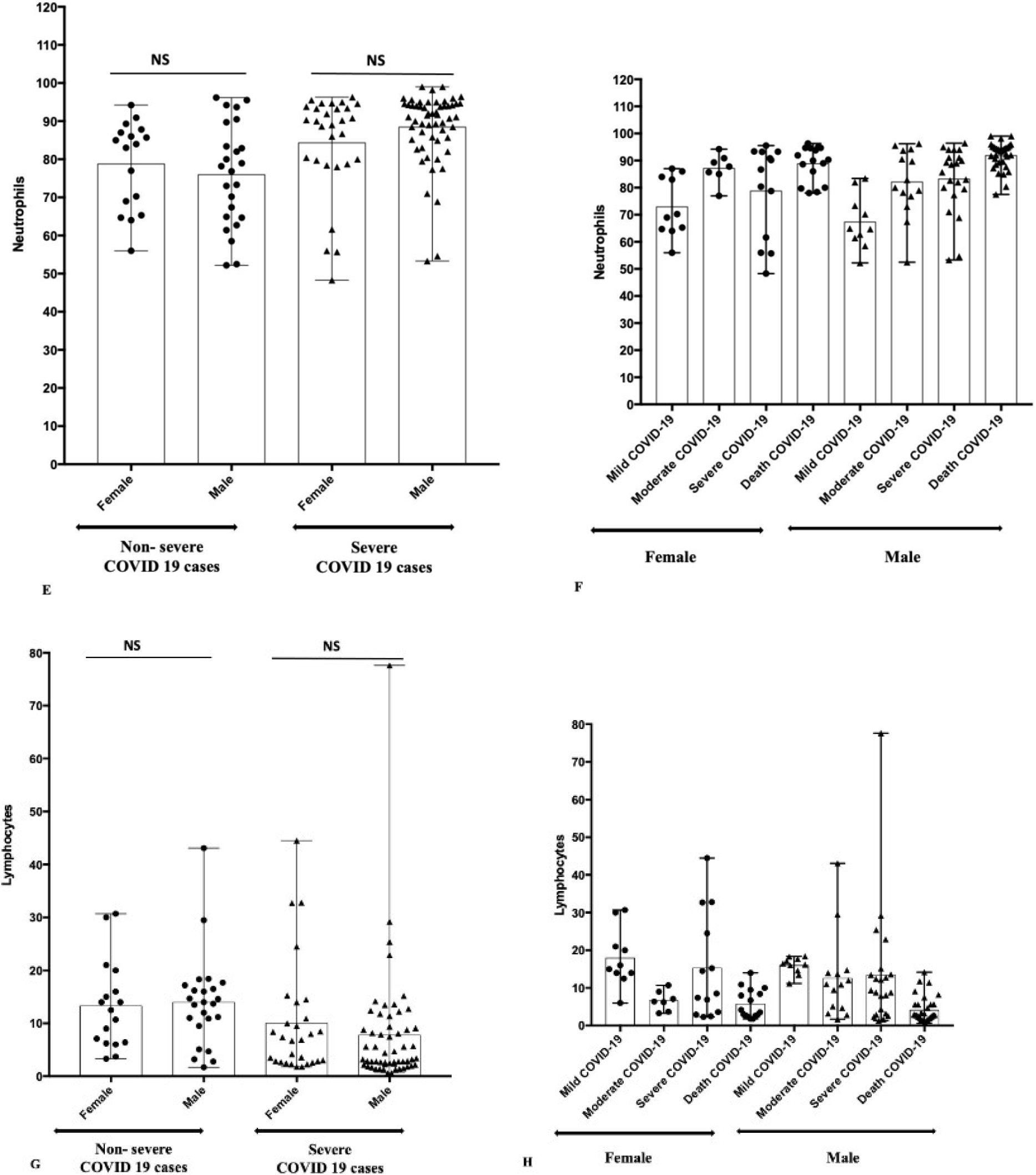
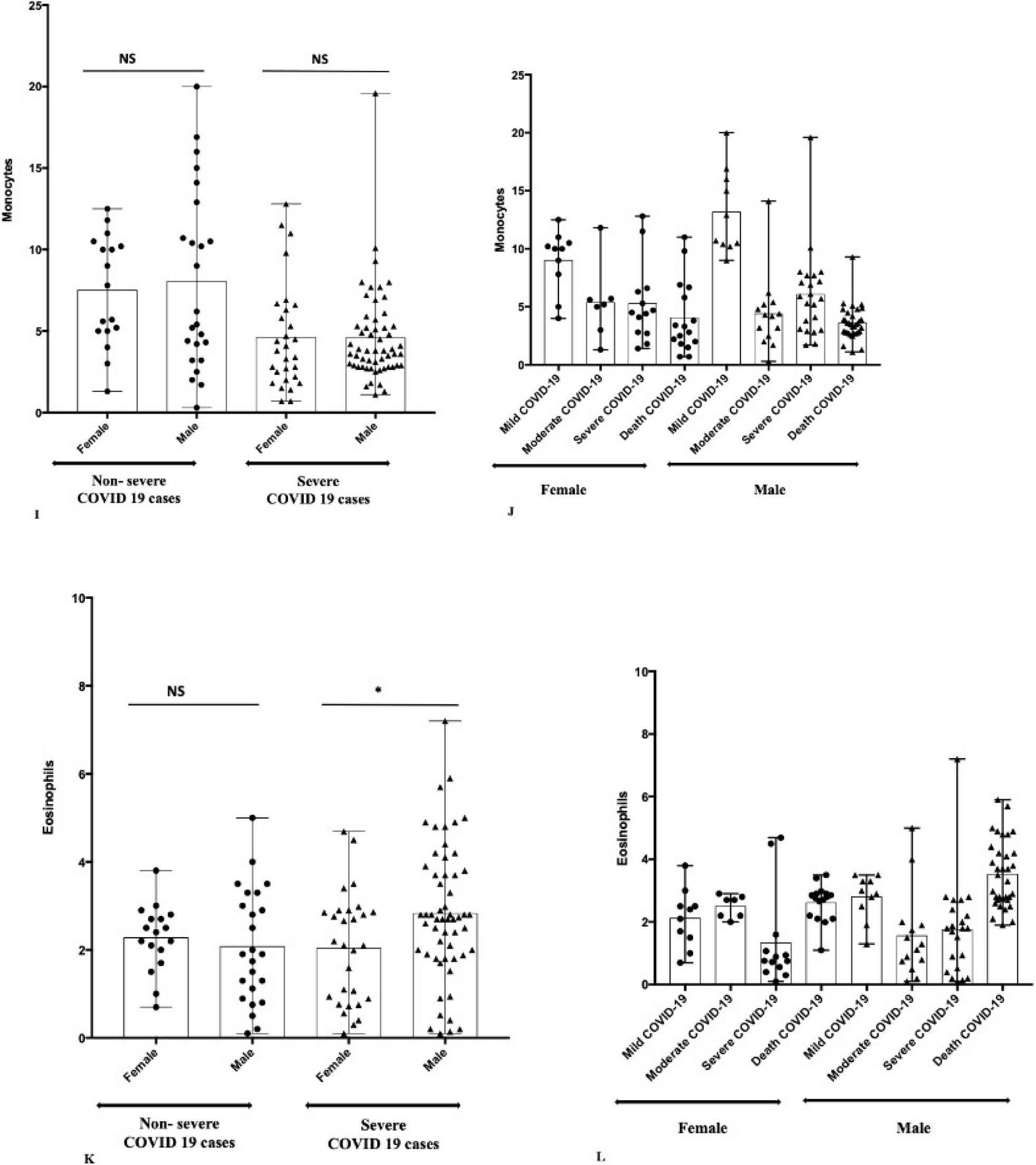
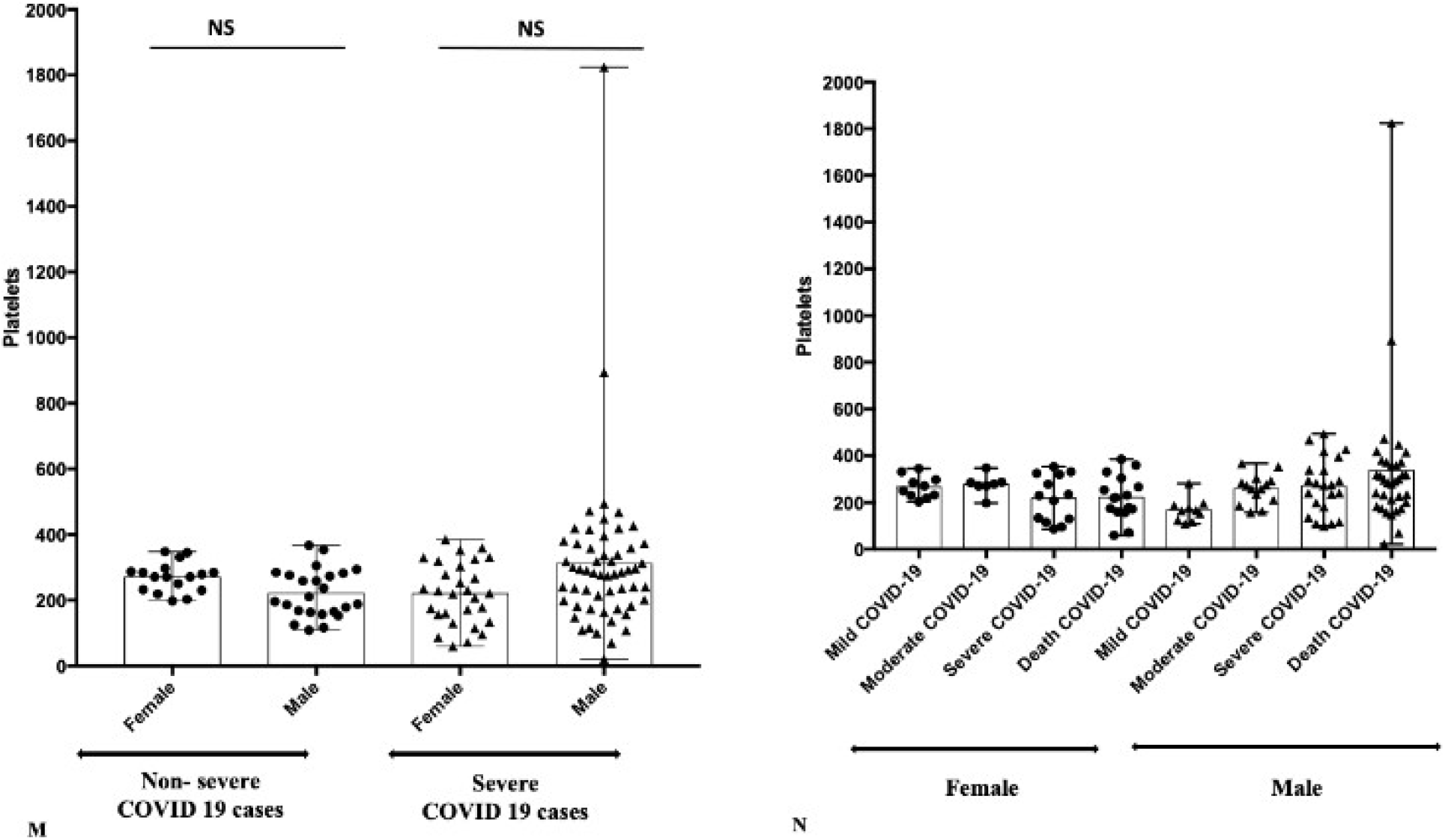
CBC of non-severe (Mild+Moderate cases) and severe COVID-19 (Severe+Death cases) Patients; (A-B) HB, (C-D) TLC, (E-F) Neutrophils, (G-H) Lymphocytes, (I-J) Monocytes, (K-L) Eosinophils, (M-N) Platelets. Figure representing the Mean±Range, * P=0.01, ** p<0.05, **** P=0.0001, NS-Non significant

#### Total Leukocyte Count (TLC)

The median TLC value was low for the non-severe group of both females and males, i.e., 10 × 10^3^/μl and 12.37 × 10^3^/μl, respectively. Similarly, the median TLC value was high for the severe group of both females and males, i.e.,12.49 × 10^3^/μl and 14.23 × 10^3^/μl. However, no significant differences were observed between males and females in the non-severe (p= 0.701) and severe (p= 0.141) cases, whereas a significant P-value was found in the case of non-severe vs. severe (p= 0.022) patients (Figure 1, Table 1). The range of variation in TLC was more in dead (3.1-32 × 10^3^/μl) COVID-19 female patients compared to critical (4.56-23.7 × 10^3^/μl), mild (5-16 × 10^3^/μl), and moderate (11.5-20.8 × 10^3^/μl) cases. The range of variation in TLC was less in mild (4.52-15.6 × 10^3^/μl) and critical (5.11-18.2 × 10^3^/μl) COVID-19 patients and a higher difference was observed in dead (3.24-40.7 × 10^3^/μl) and moderate (4.4-30.2 134 × 10^3^/μl).

#### Neutrophils

The median neutrophils were found to be low in non-severe males (77.51 %) and females (84%) compared to severe COVID-19 males (91.31%) and females (90%). No significant differences were observed between males and females in the non-severe (p= 0.451) and severe (p= 0.169) cases, whereas a significant P-value was found in the case of non-severe vs. severe (p<0.001) patients (Figure 1, Table 1). The range of variation in Neutrophils was found to be less in moderate (77-94.2%) and dead patients (78-96.3%) and high in critical (48.3-95.5%) and mild (56-87%) COVID positive females. In the case of male COVID patients, the range of variation was more in moderate (52.5-96.2%) and critical (53.3-96.4%) patients.

#### Lymphocytes

The median lymphocyte was higher in non-severe females and males, i.e., 12.5% and 13.85 %, respectively, while a low median was observed in the case of severe COVID-19 females and males, i.e., 6.9% and 3.3% respectively. The P-value between females and males for non-severe (p=0.771) and severe (p=0.114) cases were non-significant, whereas significant (p<0.001) was observed between non-severe vs. severe. (Figure 1, Table 1). In females, the range of variation in lymphocytes was noticed less in moderate (3.3-10.7%) patients compared to mild (6-30.7%), severe (2.3-44.5%), and dead (1.8-14%) COVID positive females while in males, the range of variation was less in mild (11.2-18.4%) patients, and it was noted high in critical (1.3-77.6%) patients.

#### Monocytes

The median monocyte was found to be high in non-severe females, i.e., 7.8%, and was low in non-severe males, i.e., 5.8%. The median monocyte value was the same in severe COVID-19 females and males, i.e., 3.8%. Further, no significant difference was observed in monocyte between males and females of non-severe (p=0.723) and severe (p=0.956) groups, while in non-severe vs. severe, a significant statistical p<0.001 was observed. (Figure 1, Table 1). The range of variations in critical cases of females (1.4-12.8%) and males (1.7-19.6%) were high compared to mild, moderate, and dead COVID-19 cases.

#### Eosinophils

The median eosinophils were similar in non-severe (2.4%) and severe (2.1%) females. In the case of males, the median eosinophils were found to be high in severe patients (1.9%) compared to non-severe males (1.9%). A significant difference was found in the severe group between male and female patients (p=0.019) (Figure 1, Table 1). Like monocytes, the range of variation in eosinophil count was more in the critical group of both females (0.1-4.69%) and 164 males (0.1-7.2%).

#### Platelet

The median platelet value was high in severe males, i.e., 284 × 10^3^/μl, and was low in non-severe COVID-19 males, i.e., 203.5 × 10^3^/μl. The median platelet value was low in severe females, i.e., 221.6 × 10^3^/μl, and was high in non-severe COVID-19 females, i.e., 271 × 10^3^/μl. A a significant difference was observed in platelet count in the non-severe group between males and females (p=0.018) (Figure 1, Table 1). The platelet count difference was higher in death COVID-19 cases of both females (60-385 × 10^3^/μl) and males (21-1824 × 10^3^/μl).

### Inflammatory markers Analysis in COVID-19 patients

#### D-Dimer

The median D-dimer value was 0.8 and 0.54 μg/ml in non-severe females and males. The value was high in severe female and male COVID-19 patients, i.e., 1.69 and 2.81 μg/ml, respectively. Significant P-value was found in non-severe vs. severe (p<0.001) and in severe COVID-19 patients female vs. males (p=0.002) COVID-19 patients. (Figure 2, Table 1).

**Figure 2:**
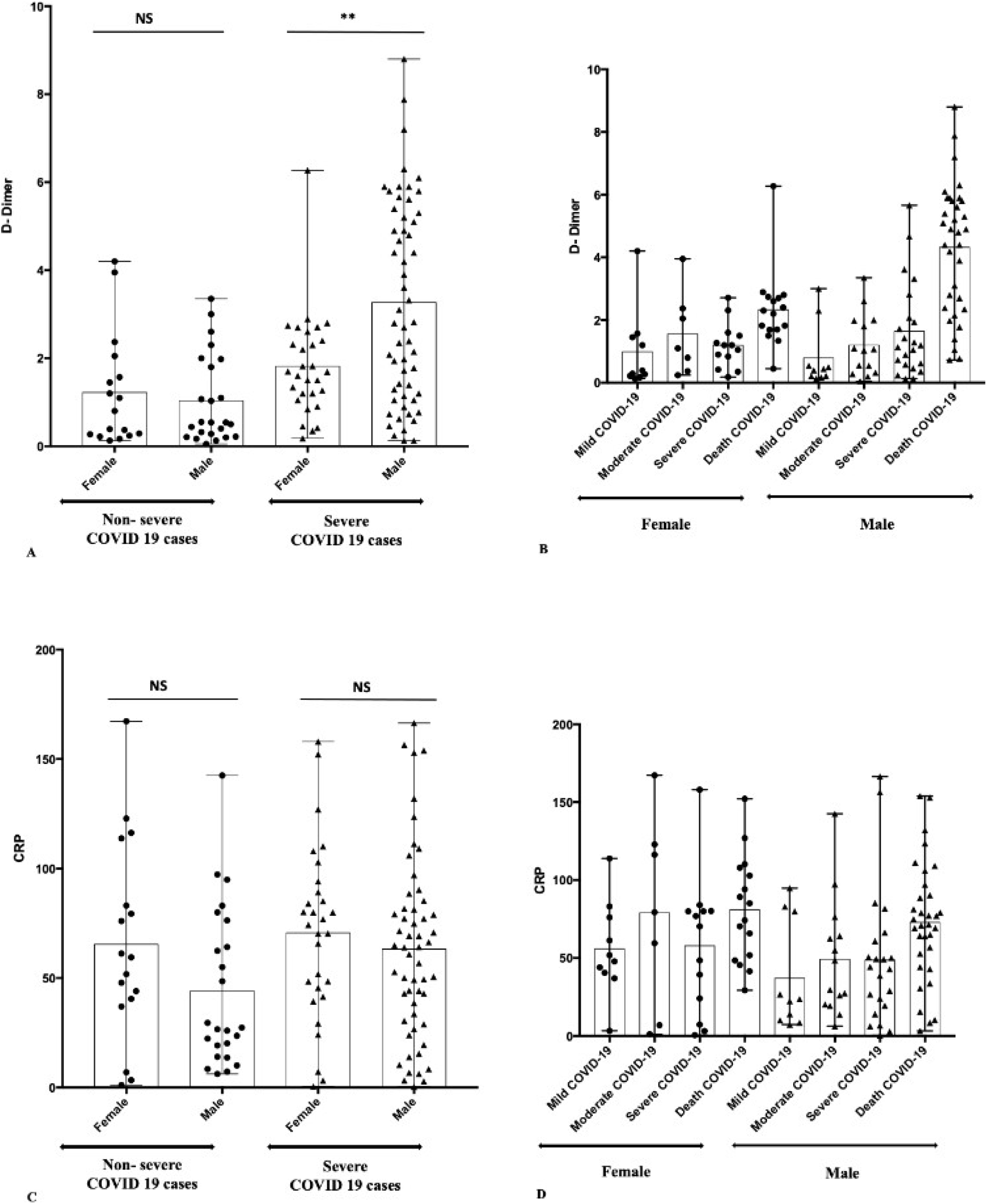
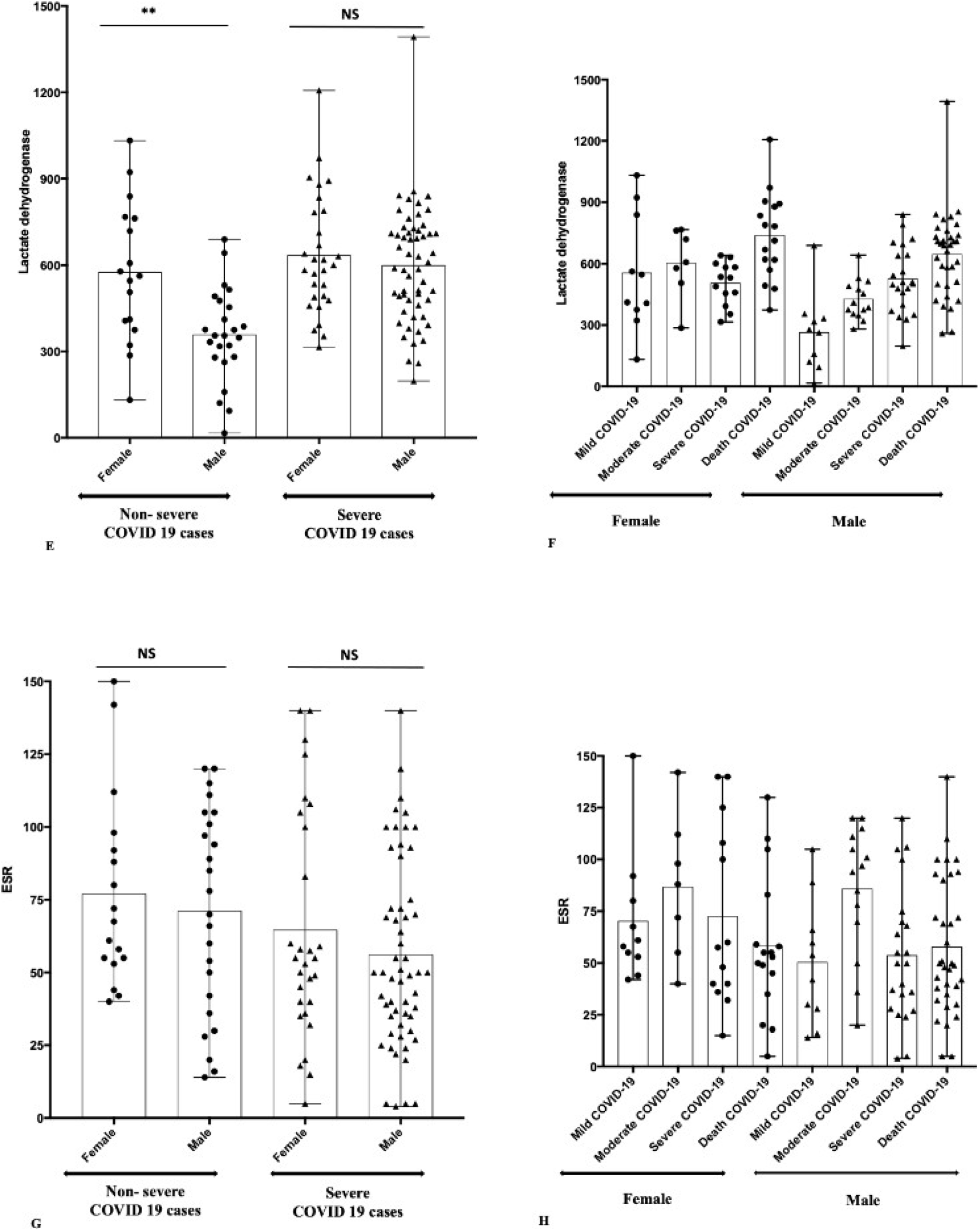
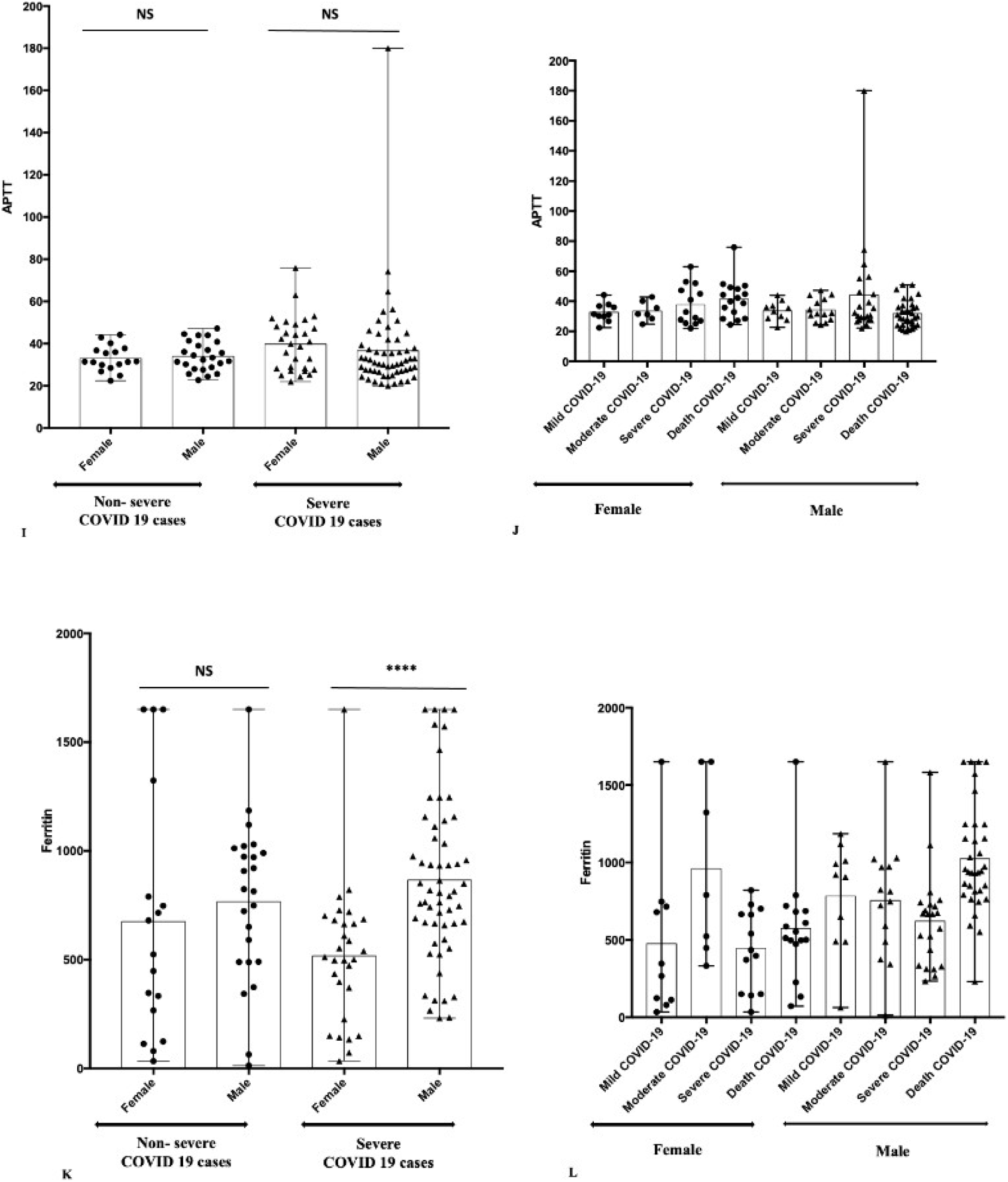
Inflammatory marker (Mild+Moderate cases) and severe COVID-19 (Severe+Death cases) Patients; (A-B) D-Dimer, (C-D) CRP, (E-F) Lactate dehydrogenase, (G-H) ESR, (I-J) APTT, (K-L) Ferritin Figure representing the Mean±Range, * P=0.01, ** p<0.05, **** P=0.0001, NS-Non significant

#### C-reactive protein (CRP)

The median CRP value was low, i.e., 27.00 mg/l in non-severe males, and was high, i.e., 64.01 mg/l in severe COVID-19 males. Similarly, the median CRP value decreased, i.e., 59.48 mg/l in non-sever females, and was high, i.e., 74.27 mg/l in severe COVID-19 females. CRP level difference was found non-significant in all the three categories studied, i.e., non-severe female vs. male (p=0.103), severe female vs. male (p=0.435), and non-severe vs. severe (p=0.104). The range of variation was less in females (0.18-2.71 mg/l) with critical symptoms and was high in dead COVID positive males (0.73-8.8 mg/l) (Figure 2, Table 1).

#### Lactate dehydrogenase (LDH)

The median was low in the non-severe group of females (562 u/l) and males (355 u/l), whereas a higher median was found in cases of females (601 u/l) and males (600 u/l). LDH values were found statistically significant in non-severe females vs. males (p=0.001) and non-severe vs. severe (p<0.001), while no significant difference was observed in the severe group of females and males (p=0.451). The range of variation was less in the female with critical (316-640 u/l) symptoms and was high in mild (132-1032 u/l) and dead (374-1207 u/l). The range of variation was less in males with moderate (281-642 u/l) symptoms and was high (259-1393 u/l) in dead COVID positive males (Figure 2, Table 1).

#### Erythrocyte sedimentation rate (ESR)

The median ESR value was high, i.e., 74 mm/hr in non-severe males, and was low, i.e., 50 mm/hr in severe COVID-19 males. Similarly, the median ESR value was high, i.e., 67.50 mm/hr in non-severe females, and was low, i.e., 55 mm/hr in severe COVID-19 females. The P-value was found non-significant in female vs. male non-severe (p=0.589) and severe (p=0.287), whereas a significant P-value was noted in non-severe vs. severe (p=0.028) COVID-19 patients. The range of variation was low in a mild case of females (42-150 mm/hr) and males (14-105 mm/hr). However, it was high in critical (15-140 mm/hr) and dead (5-130 mm/hr) females and dead (5-140 mm/hr) in male COVID-19 patients (Figure 2, Table 1).

#### Activated Partial Thromboplastin clotting Time (APTT)

The median APTT value was less, i.e., 31.45 sec in non-sever females, and was high, i.e., 32.9 sec in severe COVID-19 females. The median APTT value was a little high, i.e., 32.85 sec in non-sever males, and was low, i.e., 30.85 sec in severe COVID-19 males. No significant difference was observed in all the categories, i.e. non-severe (female vs. male; p=0.642), severe (female vs. male; p=0.498) and non-severe vs. severe (p=0.199) (Figure 2, Table 1). The range of variation was less in mild and moderate cases of COVID-19 in females and males.

#### Ferritin

The median ferritin value was similar to both non-severe (female-524 ng/ml and male 819 ng/ml) and severe (female-512 ng/ml and male 811.55 ng/ml). A significant P-value was noted in non-severe (female vs. males; p<0.001) COVID-19 patients (Figure 2, Table 1). The variation range was less in females with severe symptoms and was high in mild, moderate, and dead COVID-19 positive females. The variation range was less in males with mild symptoms and was high in moderate COVID-19 positive males.

### Kidney Function Test (KFT) markers Analysis in COVID-19 patients

Various parameters for KFT (Urea, Creatinine, Uric acid, Sodium, Potassium, and Chlorine) were assessed. In the non-severe category (females vs. males), patients showed no significant difference in any KFT markers (Table 1 and Figure 3). In the severe category (females vs. males), all the KFT markers showed no significant P-value except for creatinine (P=0.021) and uric acid (P=0.004); a a significant difference was observed. In the case of non-severe vs. severe COVID-19 patients, a a significant difference was observed in the values of urea (p=0.002), uric acid (p<0.001), sodium (p<0.001), potassium (p=0.042) and chlorine (p<0.001) whereas no difference was seen in creatinine between male and female patients(p=0.4000).

**Figure 3:**
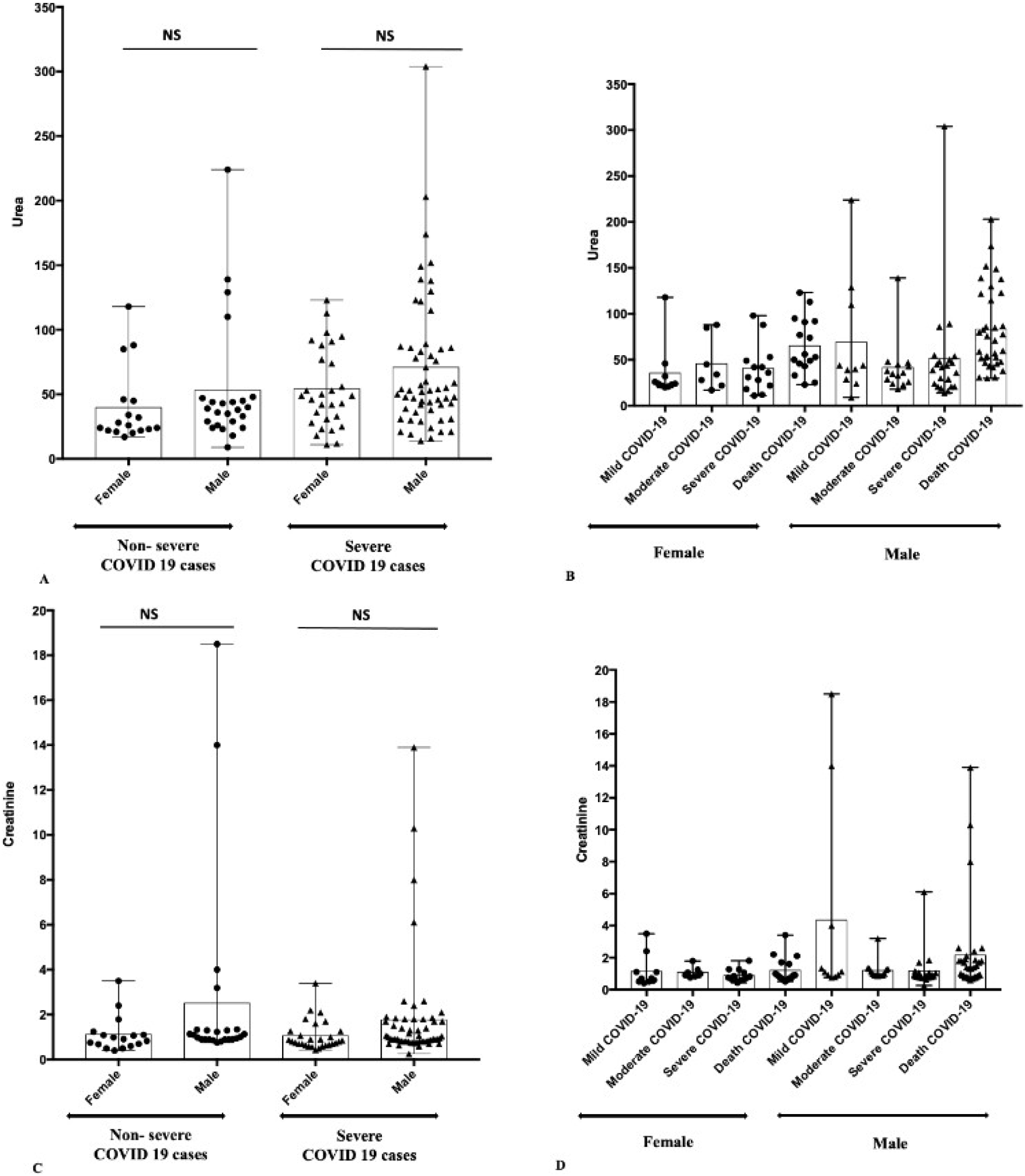
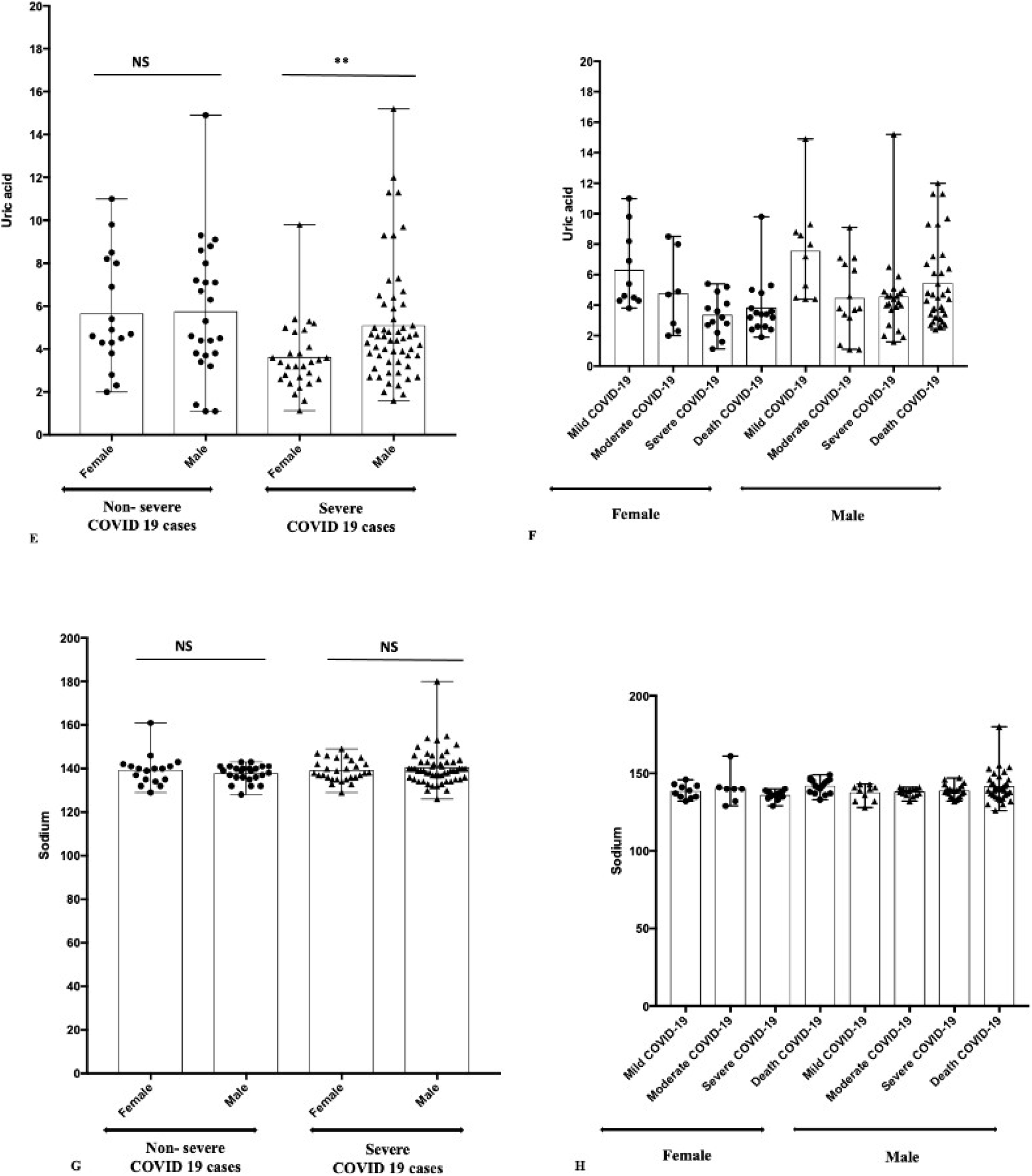
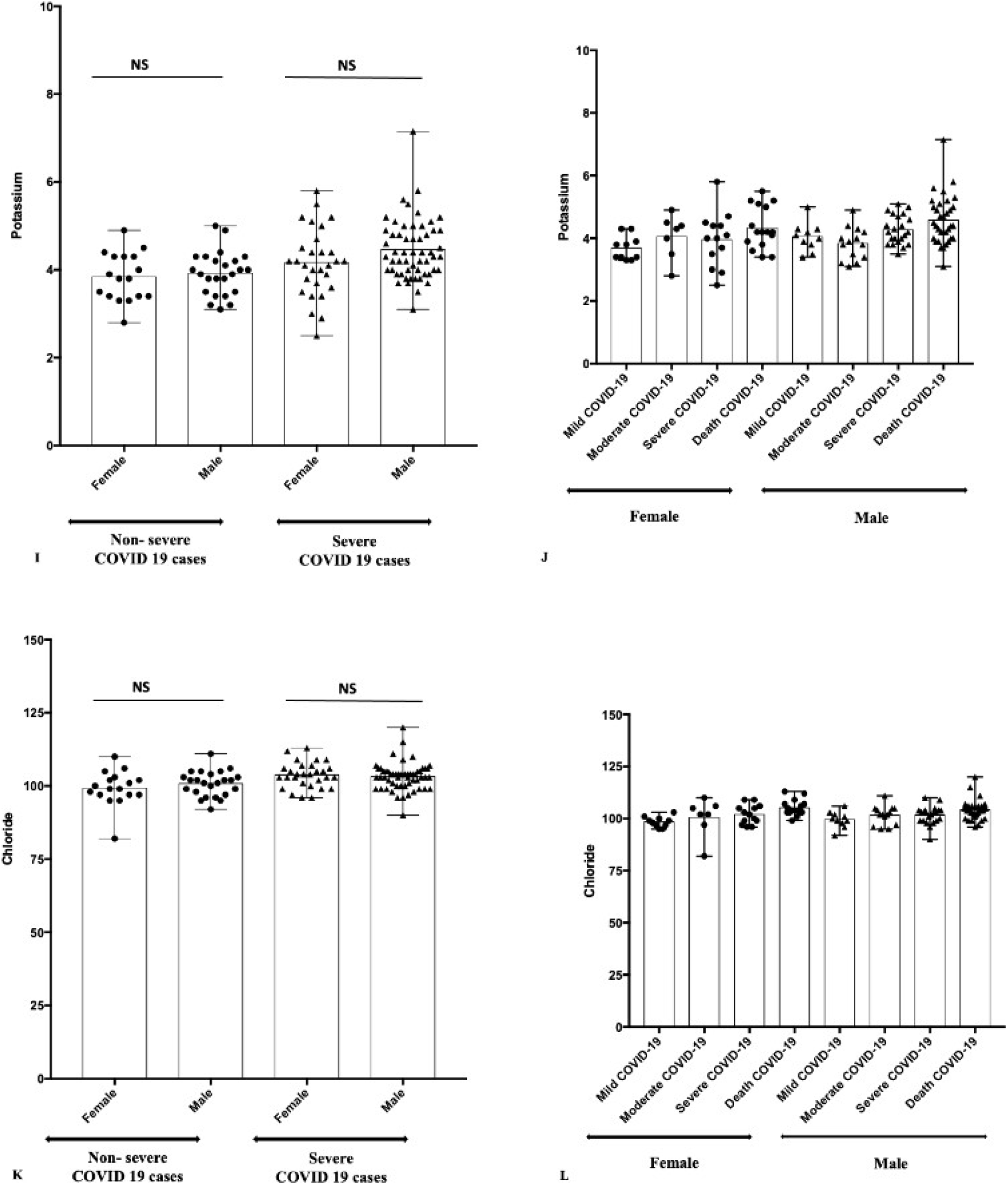
Kidney function markers (Mild+Moderate cases) and severe COVID-19 (Severe+Death cases) Patients; (A-B) Urea, (C-D) Creatinine, (E-F) Uric acid, (G-H) Sodium, (I-J) Potassium, (K-L) Chloride Figure representing the Mean±Range, * P=0.01, ** p<0.05, **** P=0.0001, NS-Non significant female patients (p=0.0059).

**Figure 4:**
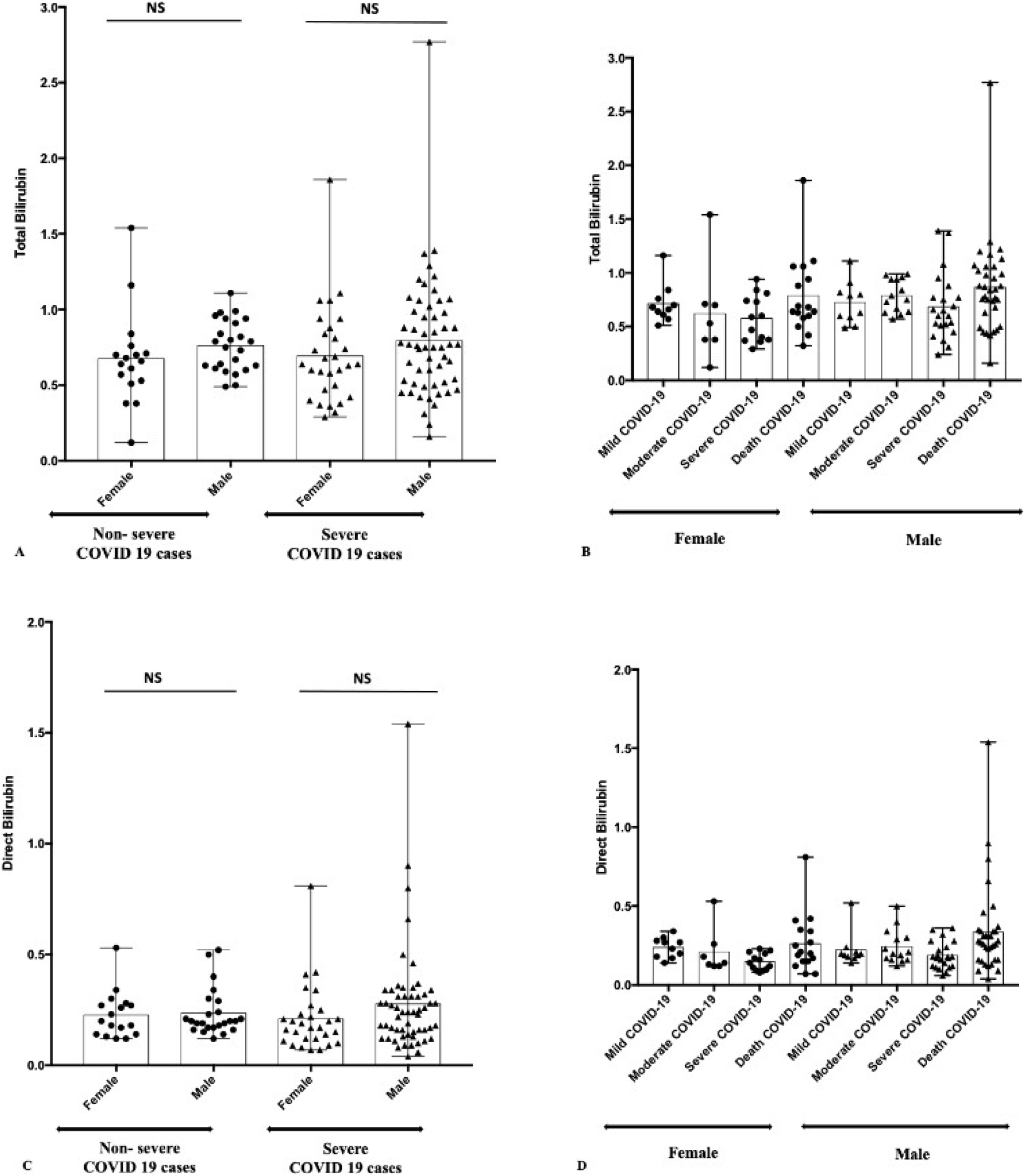
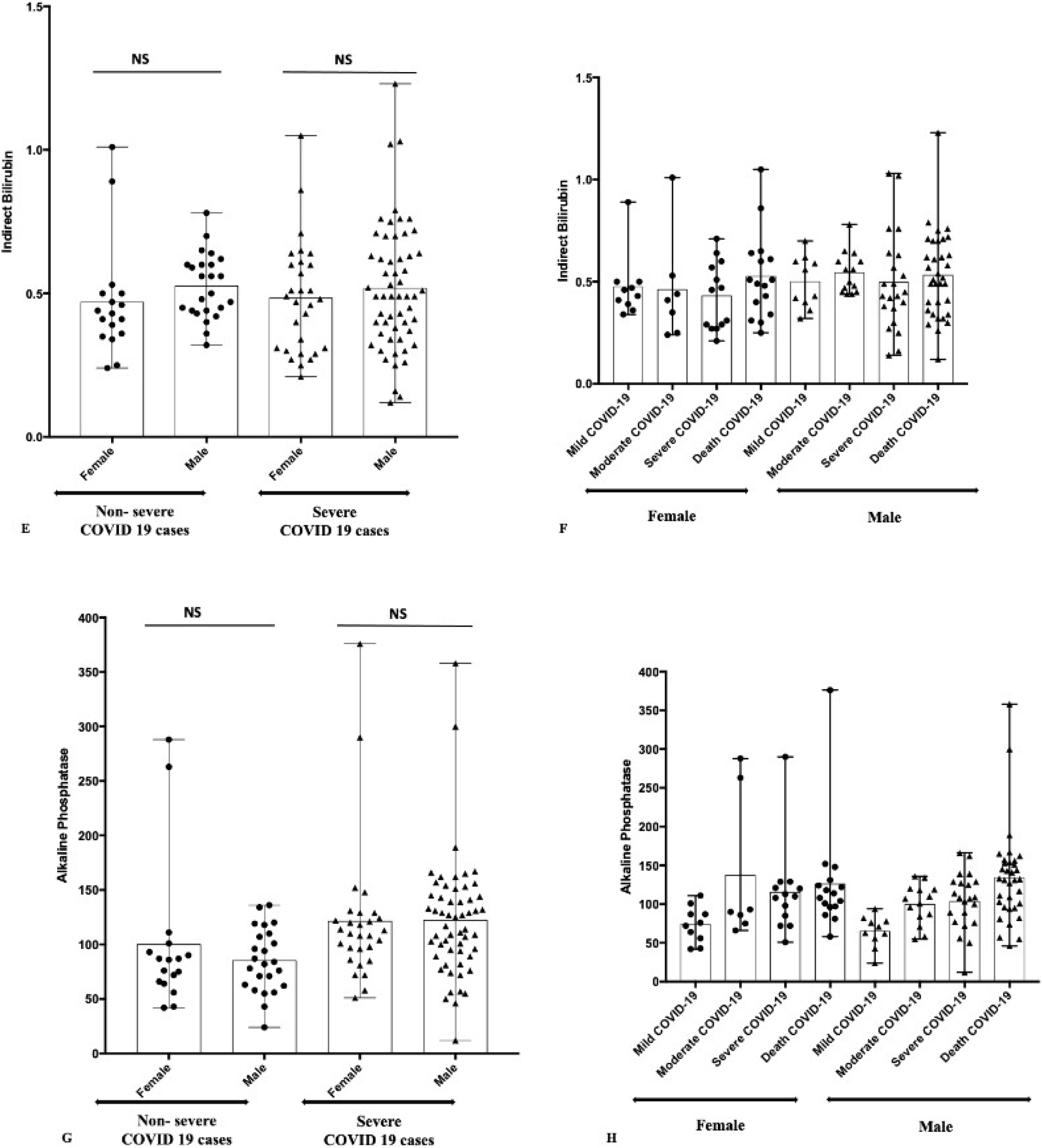
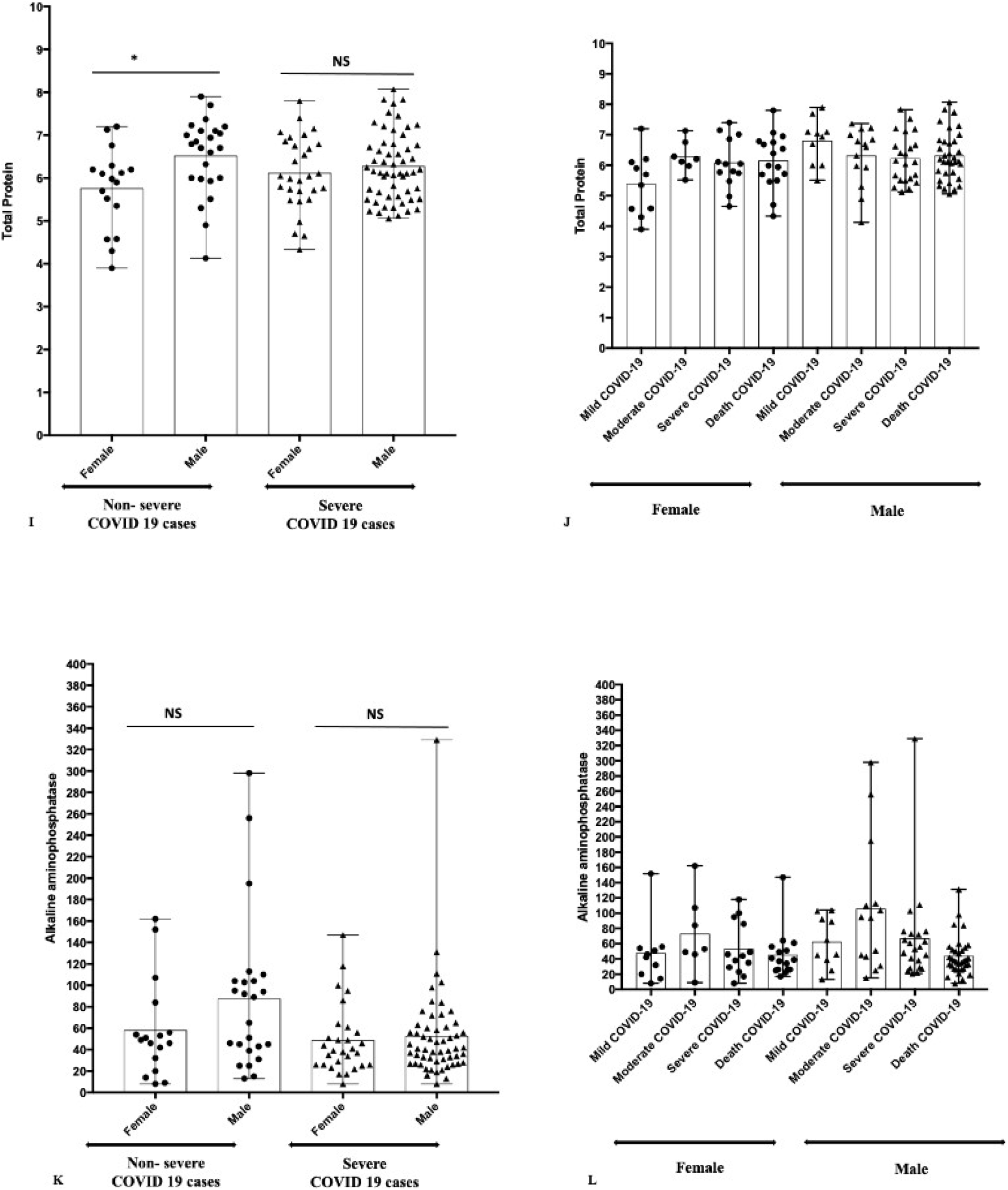
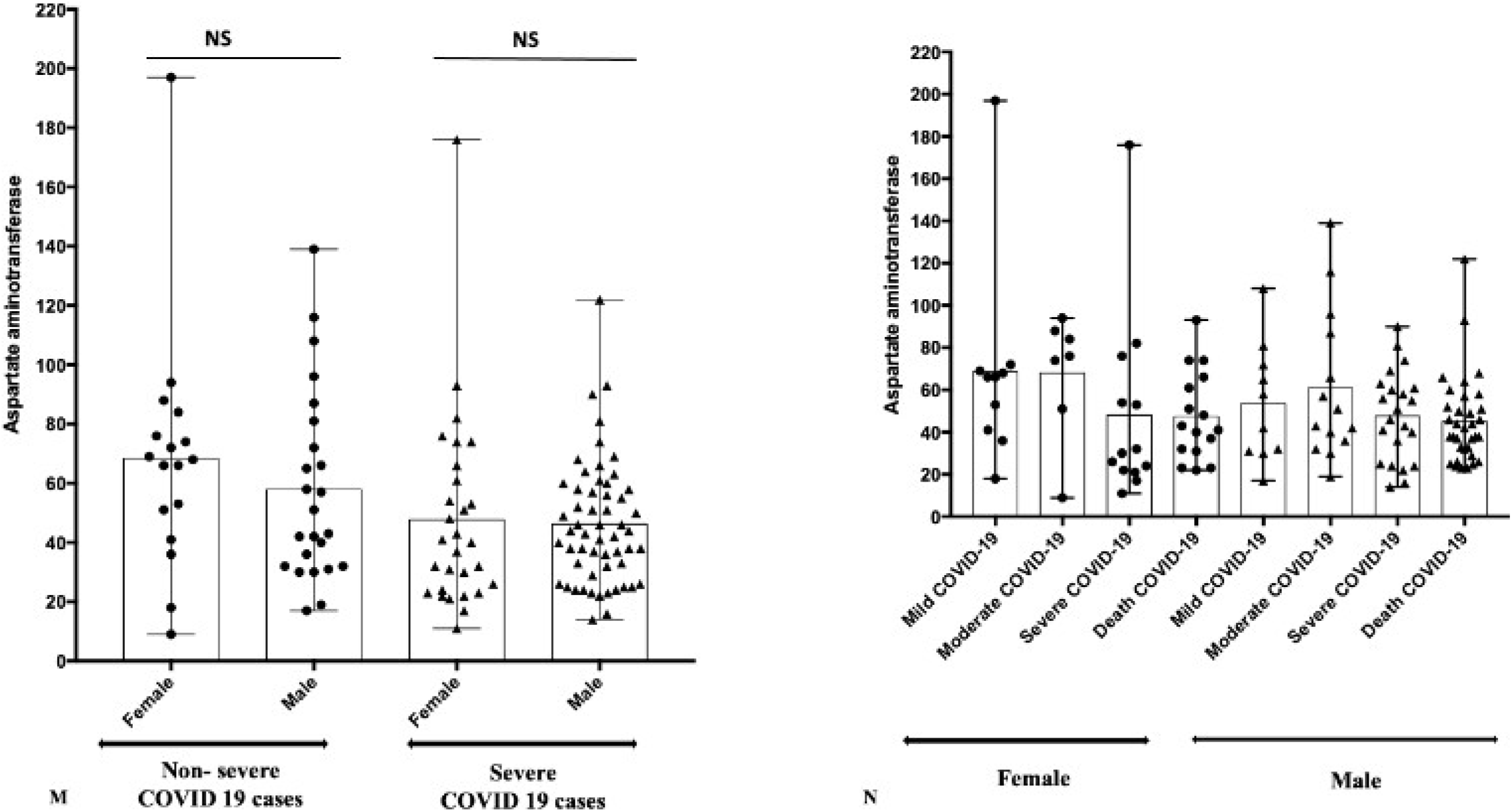
Liver function test markers (Mild+Moderate cases) and severe COVID-19 (Severe+Death cases) Patients; (A-B) Total Bilirubin, (C-D) Direct Bilirubin, (E-F) Indirect Bilirubin, (G-H) Alkaline Phosphatase, (I-J) Total Protein, (K-L) Alkaline amino phosphates, (M-N) Aspartate aminotransferase. Figure representing the Mean±Range, * P=0.01, ** p<0.05, **** P=0.0001, NS-Nonsignificant

### Liver Function Test (LFT) markers Analysis in COVID-19 patients

The LFT was evaluated by screening all included patients with mild, moderate, severe, and dead patients’ group on various parameters, namely total bilirubin direct, indirect, and alkaline phosphatase(ALP), total proteins, alanine aminotransferase (ALT), aspartate aminotransferase (AST). In the non-severe group, females vs. males showed no difference in P-value for all the parameters checked except total protein (p=0.018), found to be significant. All the checked parameters were found non-significant for severe group females vs. males. Significant P-value found for ALP (p=0.004), ALT (p=0.032) and AST (p=0.009) in non-severe vs. severe category of COVID-19 patients.

## Discussion

The variable course of illness ranges from asymptomatic to severely ill with complications of acute respiratory failure makes it crucial to collect strong evidence to determine the patient’s condition promptly and predict complications. Biomarkers are quantitative measurements that reflect the pathophysiology of disease and thus help clinicians recognize the severity of medical illness. They also aid in the developing of clinical care management algorithms that have the potential to improve patient outcomes. These laboratory indices will help differentiate severely ill patients and allow for the appropriate allocation of healthcare resources. These biomarkers in understanding COVID-19 may also help prevent the virus-induced acute inflammatory response complications such as acute hypoxemic respiratory failure and multiorgan dysfunction including acute cardiac, hepatic, and renal injury in affected patients(6).

The increased total leukocyte count is an indicative marker for detecting an infectious condition in most diseases.(10)Thus, previous studies also evaluated leucocytes’ role in the COVID-19 condition. (14,15,22)In their retrospective study, Qin et al. concluded that severe patients showed leukocytosis compared to non-severe COVID-19 patients. In our present study, we also found leukocytosis in severe cases, and the result was comparable with various other studies. (15,22)

The NLR, calculated simply by the ratio of neutrophils count/lymphocytes count, is an an inflammatory marker that can predict the probability of death in patients with various cardiovascular diseases (23,24). Moreover, NLR has been identified in a meta-analysis as a prognostic biomarker for patients with sepsis(25). For COVID-19 patients, NLR has been shown to be an independent risk factor for severe disease (26–28). Fifty (75.8%) patients with disease progression during hospitalization had an NLR of 2.973 (29), which may indicate COVID-19 infection severity (30). The binary logistic analysis identified elevated NLR (hazard risk (HR): 2.46, 95% confidence interval (CI): 1.98–4.57) as an independent factor for poor COVID-19 clinical outcome (31), which was confirmed by a meta-analysis that reported that NLR values were significantly increased in severe COVID-19 patients(32). NLR elevation maybe due to dysregulated expression of inflammatory cytokines, an aberrant increase of pathological low-density neutrophil and the upregulation of genes involved in lymphocyte cell death pathway, caused by the mechanism of SARS-CoV-2 infection (33). In our study too an elevated NLR was observed corroborative with published literature. Our analysis also indicates that low neutrophil and lymphocyte counts showed severe COVID-19 and were analogous to the previous findings (10,11). Infection, (33)Lymphopenia occurs due to excessive activation of the inflammatory cascade and cardiac involvement is a crucial feature of COVID-19 disease and has a high predictive value. However, the understanding of the underlying mechanisms is still limited(34). Based on the observations derived from clinical practice, it has also been postulated that coronaviruses may directly infect bone marrow precursors, resulting in abnormal hematopoiesis, or trigger an auto-immune response against blood cells (35,36). In severe disease, WBCs show lymphocytopenia, affecting CD4. and CD8. Cells, a decrease in monocytes and eosinophils, and a clear increase in neutrophils and NLR. These simple parameters might use for early diagnosis and identification of critically ill patients(37,38). On the other hand, similar to our finding, Pan et al. also showed no variation in monocyte count in COVID-19 patients(39). In contrast with Pan et al., the present study showed no significant increase in Hb observed in the case of severe COVID-19 patients (39).

As platelet count is a simple, cheap, and easily available biomarker and has been independently associated with disease severity and mortality risk in intensive care unit (ICU) H5 (40–42), it has been rapidly adopted as a potential biomarker for COVID-19 patients. The number of platelets was reported to be significantly reduced in COVID-19 patients (43,44) and was lower in non-survivor patients compared to survivors. (45) Low platelet count has been associated with increased risk of severe disease and mortality for COVID-19 patients, and can serve as an indicator of clinical disease worsening during hospitalization. (46) Another research the group found that patients with severe pneumonia induced by SARS-CoV2 had higher platelet count than those induced by non-SARS-CoV2. (47)The patients with significantly elevated platelets and higher platelet-to lymphocyte ratio (PLR) during treatment had a longer average hospitalization days. (48) Damaged lung tissue and pulmonary endothelial cells may activate platelets in the lungs, resulting in the aggregation and formation of microthrombi, thereby increasing platelet consumption. (49) The previous meta-analysis reported that platelets were not an appropriate marker for considering the severity of COVID-19 disease.(13,14) Similarly to this finding, our data showed no significant variation between the non-severe and severe cases. In In contrast, low platelet count was suggested in the severe COVID-19 cases. (12)

The increased inflammatory response in COVID-19 and hypoxia due to severe pneumonia eventually leads to the activation of coagulation and fibrinolysis, followed by a hypercoagulable state causing DIC and multi-organ dysfunction. (50,51)Additionally, prior studies show that D-dimer levels greater than 2.0 µg/mL on admission could effectively predict in-hospital mortality rates of patients with COVID-19.(51) Patients with higher D-dimer levels requiring intubation were also associated with a greater probability of developing a pulmonary embolism after admission. A study by Yu et al. found significantly elevated levels of D-dimer in COVID-19 patients than in patients with community-acquired pneumonia. (52) Our study also revealed that the D-dimer levels were high in severe COVID-19 cases of both male and female sex.

C-reactive protein (CRP) is a sensitive biomarker for inflammation. As an acute phase inflammatory mediator, CRP is synthesized and released by the liver into the bloodstream, contributing to the host’s resistance to invading pathogens. (1) CRP elevation may also be caused by bacterial coinfection that occurs in severe COVID-19. Moreover, a robust inflammatory the response that occurs in severe disease may cause CRP levels to increase significantly. (1) Elevated CRP levels are directly correlated with inflammation and disease severity. Hence, it is a crucial biomarker in diagnosis and assessing the severity of infectious diseases H4. In the the present study, CRP levels were significantly elevated in severe COVID-19 cases of males and females. In non-severe cases, though CRP was raised, it was not that significant.

LDH is present in tissues throughout the body and is involved in the interconversion between pyruvate and lactate through a nicotinamide adenine dinucleotide (NADH)-dependent reaction. Abnormal LDH levels can result from decreased oxygenation, leading to an upregulation of the glycolytic pathway and from multiple organ injuries. The mechanism through which lactate leads to injury are via the action of metalloproteinases and enhanced macrophage-mediated angiogenesis. (53)In a study conducted on COVID-19 patients, it was found that levels of LDH early on in the course of the disease can be a good predictor of lung injury and severe COVID-19 cases. (54) High LDH has also been associated with worse outcomes in several studies. (55–57)

In a pooled analysis, elevated LDH values were associated with a >16-fold increase in the odds of mortality and a sixfold increased odds of severe disease. (53) Our overall results demonstrated that the LDH levels were raised in non-severe COVID-19 cases as compared to severe COVID-19 cases that are contrary to the published literature.

The immunological biomarkers of serum ferritin are reported to be significantly increased in non-survivors vs. survivors (WMD: 4.6 pg/mL and 760.2 ng/mL, respectively) and as compared to severe vs. non-severe disease (WMD: 1.7 pg/mL and 408.3 ng/mL, respectively). (58) In In the present study ferritin was raised in non-severe cases of COVID-19 in both male and female whereas the rise was not that significant in severe COVID-19 cases. Similar to Izcovich et.al, we also found a significant association between non-severe and severe cases. (16)

The abnormal change in biomarkers level suggests that COVID-19 indicates various indicators of analogous organ damage. A large number of COVID-19 samples were evaluated by Guan et al. and showed an increased level of ALT, AST, and creatinine in severe cases compared to non-severe (Guan et al., 2020). Our study also showed the elevated levels of these parameters in COVID-19 cases.

## Conclusion

Various biomarkers values differing between mild/moderate versus severe and fatal cases may help in the early management of the high-risk COVID-19 patients and potentially improve prognosis and mortality rates. Furthermore, these biomarkers may help develop prevention policies and responses to combat critical adverse COVID-19 outcomes.

## Data Availability

All data produced in the present work are contained in the manuscript

